# A transparent EWMA-based early-warning index for RSV and influenza A from WastewaterSCAN wastewater signals, benchmarked against CDC laboratory surveillance

**DOI:** 10.64898/2026.01.20.26344452

**Authors:** Taylan Demir, Hasan Hüseyin Tosunoğlu

## Abstract

Wastewater based epidemiology offers a valuable population level signal for monitoring respiratory virus activity, but its routine use in public health practice requires alerting methods that are transparent, interpretable, and comparable across locations. In this study, we propose a simple early warning framework that transforms wastewater viral RNA measurements into actionable alerts using a standardized statistical process control approach. The method relies on variance stabilization, site specific baseline normalization, and an exponentially weighted moving average to identify sustained increases in viral activity. To support operational relevance, wastewater derived alerts are benchmarked against established laboratory surveillance systems using a harmonized onset definition. The proposed framework emphasizes clarity, auditability and adaptability rather than complex forecasting, enabling straightforward interpretation by public health practitioners. Our results demonstrate that wastewater signals can provide timely situational awareness for respiratory virus circulation and support their use as a complementary tool for public health surveillance and preparedness.

## 1 Introduction

Annually, seasonal respiratory viruses such as Influenza A Virus (IAV) and Respiratory Syncytial Virus (RSV) impose a great deal of stress upon the healthcare system. Seasonal fluctuations in the transmission of these viruses can lead to rapid increases in outpatient demand, hospital admissions and emergency department visits from one season to another and from year to year, resulting in additional strain on the healthcare system. Expectations for IAV and RSV activity require timely situational awareness of not only the clinically significant aspects of IAV and RSV activity, but also the operational demands placed on the public health agency and the health authorities. By having sufficient awareness of IAV and RSV activity, public health agencies will be able to coordinate their messaging, develop consistent case definitions and manage their capacity to respond to an IAV and/or RSV surge more effectively [1–5]. Wastewater-based epidemiology (WBE) offers a complementary signal for surveillance, as it compiles biological material removed by the inhabitants of an entire city or region and reduces bias resulting from how people seek healthcare or obtain tests through traditional means. Rapid collection of WBE values allows for timely input for public health decisions rather than relying solely on retrospective studies [6]. However, raw WBE values do not always provide an easy to interpret early warning sign with regards to infection, disease transmission, or potential risks. For example, wastewater contains inhibitors as well as diverse backgrounds of matrix materials (organic mater and salts) that can complicate the analytical method and have major influences on the time of signal appearance (alert). A common theme in the methodological literature is the importance of accuracy, timeliness for operational use, and comparability across multiple jurisdictions [8]. In the case of influenza A, it has been demonstrated by multiple studies that the RNA signals in wastewater, especially with respect to settled solids, track clinical dynamics and epidemiological patterns. Specifically, in a large prospective study, the IAV RNA concentration values measured in solids collected from various municipal wastewater treatment facilities tracked closely with hospitalizations and influenza-like illnesses’ information regarding onset, offset, and peak dates [8]. The research also demonstrated the inherent variability of methodologies for establishing a baseline, which is vital for making timely public health decisions, based upon the aforementioned correlation between hospitalization and documented evidence of the epidemic. In summary, the field of using wastewater to assess the presence of viral activity is now evolving from examining the existence of viral load within wastewater to the development of an acceptable practice for converting a continuous signal into a stable, operational “trigger” for epidemic preparedness. Although there has been a rapid increase in the amount of published evidence that points to the promise of monitoring wastewater for RSV, the results of various studies on this topic reflect the need for standardized alerting definitions. A large, multi-site study in the U.S. indicated that concentrations of RSV RNA detected in wastewater solids can vary widely and also correlate to broader indicators of RSV burden in the clinical setting [10]. It should be noted, however, that studies comparing cross-jurisdictional data illustrate that it is inappropriate to create “one-size-fits-all” rules for determining the start of RSV based on wastewater concentrations; in some jurisdictions, there are earlier observed starts of RSV than the corresponding clinical diagnoses, while others show that clinical diagnoses preceded the collection of wastewater [10]. The differences observed may be due to several factors, including testing protocols, delays in reporting, differences between the areas sampled, and biological or behavioural differences affecting virus shedding and transport in the wastewater. Instead of interpreting these discrepancies to suggest that wastewater predictive rules are not reliable, one could view the differences between jurisdictions as information suggesting that alerting procedures for wastewater monitoring should be developed and validated against the operational goals for wastewater monitoring, such as increasing the positive predictive value of wastewater for indicating true clinical burden and reducing the number of false alerts during off-seasons [11,12]. Surveillance users continue to experience the practical challenge of creating an effective early-warning index due to advances made with regard to measurement and validation. One huge component of this challenge is that we need an easy-to-read, defensible method for creating a reproducible early-warning index that can be benchmarked against results produced via traditional laboratory surveillance. The majority of existing approaches for early warning indexes can be classified as using overly-complicated models or ad hoc thresholds, neither of which lends themselves to easy communication and auditing [13,14]. By using the statistical process control method as a basis for detecting and managing persistent shifts occurring within varying degrees of noise, with respect to measured parameters of interest, we are able to use very clear and defined rules regarding how to interpret, audit, and stress-test the alarm threshold to get to the point of not reacting to a false alarm. The EWMA method or chart is simple and easy to understand, and has been around for a long time and widely studied. We establish an Early Warning Index (EWI) using WastewaterSCAN Settled Solids RNA measurements for Respiratory Syncytial Virus (RSV) and Influenza A. Our intention is not to create a complex forecasting system, but rather to develop a simple, reproducible procedure that can convert standardized wastewater signals into easy-to-read warning alerts and allow for equitable benchmarking of these alerts against typical laboratory surveillance systems. In addition to developing the EWI, we fully describe the methods used to develop the EWI and assess its operational efficiency. This includes (i) variance stabilization transformation and standardizing to seasonal low-activity levels, (ii) creating an EWI alerting system based upon an EWMA system with defined transparent parameters and persistence requirement, and (iii) measuring EWI performance utilizing lead time, false alerts during non-influenza seasons, and sensitive testing of candidate EWI parameters. Each of these performance criteria bridges the connection between wastewater-derived real-time data and public health notification systems, allowing for accurate evaluation of other methodologies.

## 2 Data

### 2.1 Wastewater RNA signals (WastewaterSCAN)

We conducted an analysis of site-specific time series obtained from 24-hour composites of wastewater settled solids for RSV and Influenza A RNA detected by WastewaterSCAN at 92 number of facilities located across regions defined within the study period (epidemiological week 35/2022 through week 18/2025). Settled solids from the wastewater offer an enriched and relatively stable matrix for monitoring respiratory viruses. Normally the detection of RSV and Influenza A RNA in the settled solids is much better and more stable than what would be expected from liquid wastewater and will therefore allow high-frequency sampling to take place. Therefore, monitoring settled solids provides an opportunity to monitor respiratory viruses at very high frequencies with the potential to trigger alerts to the community on a weekly basis without the need for modelling and complex statistical analyses. Using droplet digital RT-PCR (ddRT-PCR), WastewaterSCAN quantifies RNA from the Wastewater (settled solids) of each of the viral types (RSV and Influenza A) sampled, and samples are always analysed in duplicate using standardized laboratory protocols to ensure reproducibility.

#### Wastewater measurement definition

For each site *s*, virus *v* ∈ {RSV, IAV}, and epidemiological week *t*, we denote by *x*_*s,v,t*_ the reported concentration of viral RNA in settled solids (units as reported by WastewaterSCAN).

#### Assay targets and detection limits

Assay targets, limits of detection (LOD), and limits of quantification (LOQ) followed the standardized WastewaterSCAN laboratory protocol and reporting conventions. Non-detects were retained as reported by WastewaterSCAN; when only a non-detect flag was available, values were set to 0 prior to the log_10_(1+*x*) transformation.

Research has demonstrated that RT-PCR of settled solids will reflect accurately the dynamics within the community for a number of different viral diseases [6,8]. Logged concentrations of wastewater RNA from settled solids were transformed using a variance-stabilising logarithm and standardised for every sampling site according to the baseline activity levels for low sampling volumes (see Methods). The WastewaterSCAN “input database” consists of the average weekly data collected for all WastewaterSCAN sites from the entire WastewaterSCAN surveillance network over the last 12 months. The aggregated data was used to provide a benchmark for future monitoring efforts against clinical data [6,8].

### 2.2 RSV laboratory surveillance (NREVSS)

In order to evaluate alerts based on wastewater, we have established a clinical benchmark for comparison by utilizing weekly surveillance indicators of RSV from the National Respiratory and Enteric Virus Surveillance System (NREVSS) in the U.S. Participation in the NREVSS is voluntary and therefore each week clinical and public health laboratories report their week’s testing volume (i.e., Tested Count) and number of positive tests (ie., Positive Count). Using this information we derive laboratory positivity metrics including percentage of positive to tested counts and weekly positive testing counts from data reported by the NREVSS [15]. To allow for a valid comparison between wastewater-based alerting systems and laboratory-defined onsets of disease, we align the NREVSS series to epidemiological weeks and apply the same persistence criteria defined by the wastewater alerting framework in defining both onset and transition to the alert phase. The weekly volume of tested RSV positive tests provided by NREVSS will be utilized to generate the series which forms the basis of our laboratory benchmark for onset and performance evaluation [15].

### 2.3 Influenza A laboratory surveillance (U.S. virologic surveillance reported via FluView streams)

In the current study, influenza A metrics have been determined using CDC FluView and FluView Interactive virologic surveillance data.Weekly virologic surveillance data reported through CDC FluView/FluView Interactive are utilized to create Influenza A benchmarking in this study. These data sets summarize the volume of laboratory testing (total testing volumes) and the number of positive laboratory results (total positive laboratory results) throughout the laboratory networks in the U.S. The benchmarking also provides a means to construct positive laboratory result counts and percentage positive laboratory results for Influenza A [16]. Additionally, in addition to reporting overall numbers of positive and negative laboratory results, virologic surveillance also allows weekly breakdowns of laboratory results by specimen category (and, if available) by influenza types/subtypes, enabling for consistent tracking of overall activity of Influenza A [16]. In order to provide a basis for the comparison of wastewater-based alerts with laboratory alerts, we aligned our laboratory series to epidemiologically defined weeks and will determine at what point (alert/onset) each emission led to a laboratory tests being positive (see methods). This link allows us to provide a common format for quantifying how frequently wastewater alerts come before laboratory-confirmed increases in influenza A activity, and also how frequently alerts are sent during off-seasons.

### 2.4 Time alignment and basic quality control

The wastewater measurements were aligned with the laboratory surveillance stream on a common time base of epidemiological week to eliminate bias caused by unequal reporting frequencies between the two sources of data. In cases where multiple wastewater measurements from the same site occurred during the same epidemiological week, we summarized that week using the weekly median to obtain a single weekly value. This weekly aggregated value corresponds with routine public health reporting and allows for a consistent implementation of the alert criteria [6,8]. To effectively maintain transparency and mitigate the risk of making unfounded assumptions that may unduly influence the timing of alerts, we took steps to implement basic quality control procedures. We implemented basic quality control procedures through the following actions: First, we required that each site maintain a consistent and non-duplicated week index (or timeline); Second, we established an explicit record for weeks that were missing; Third, we screened for obvious discrepancies in processing. We did this by identifying cases where there were large jumps in values from one week to the next or by comparing the week-by-week values with both laboratory/quality control (*QC*) notes and the expected epidemiological values. If any weeks were flagged, we handled these weeks conservatively (see Methods) and performed sensitivity analyses to understand the extent to which they could influence alert timing.

## 3 Methods

### 3.1 Weekly aggregation and notation

For each wastewater site *s* and virus *v* ∈ {RSV, IAV}, let *x*_*s,v,t*_ denote the reported concentration of viral RNA in wastewater settled solids at epidemiological week *t*. Weekly aggregated levels from wastewater analysis are created by combining all measurements taken during a week into one robust summary (weekly median),

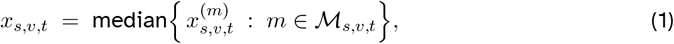

where 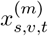 denotes the *m*-th measurement collected for site *s* and virus *v* during week *t*, and ℳ_*s,v,t*_ indexes all measurements available for (*s, v, t*). This method is used for comparison purposes between the weekly aggregated summaries of wastewater analyses at the national level and weekly clinical indicators [8].

In laboratory surveillance, we construct a weekly comparator series *y*_*r,v,t*_ for each reference area *r* (e.g., a U.S. state or HHS region), virus *v* ∈ {RSV, IAV}, and epidemiological week *t*. In this study, we take *y*_*r,v,t*_ to be the weekly count of positive laboratory tests reported by the relevant CDC surveillance stream (NREVSS for RSV; FluView/FluView Interactive for influenza A). Each wastewater site *s* is mapped to a reference area via *r* = *r*(*s*), so that site-level wastewater alerts can be evaluated against the corresponding laboratory activity series *y*_*r*(*s*),*v,t*_.

Matched laboratory preprocessing and onset. To enable a direct, apples-to-apples timing comparison, we apply the same variance-stabilization, standardization, smoothing, thresholding, and persistence logic to the laboratory series as used for wastewater. Define the transformed laboratory series

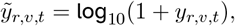

and let *T*_*r,v*_ be the set of epidemiological weeks with an observed laboratory value for (*r, v*). Analogous to (3), define baseline weeks

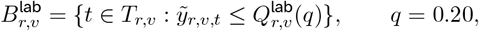

where 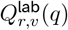 is the empirical *q*-quantile of 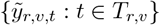. Compute baseline moments

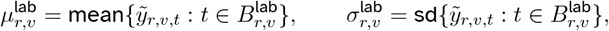

and form the standardized laboratory series

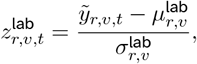

with the convention 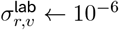 if 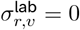.

We then compute a laboratory EWMA score using the same smoothing parameter *λ*:

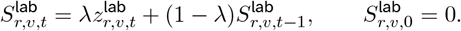

If a laboratory value is missing at week *t*, we carry forward 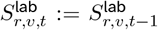. Finally, fixing the same threshold *h* and persistence *k* as in (8)–(9), define

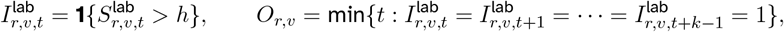

with *O*_*r,v*_ = ∞ if no such run occurs. The laboratory-defined “in-season” weeks are taken to be the union of weeks in laboratory alert episodes (defined analogously to Section 3.4) for (*r, v*).

### 3.2 Variance stabilization and baseline standardization

Wastewater RNA concentrations are typically right-skewed and can vary over several orders of magnitude. To stabilise variance and reduce the influence of extreme values, we apply a log transformation to the weekly aggregated concentrations. For each site *s* and virus *v* ∈ {RSV, IAV}, define

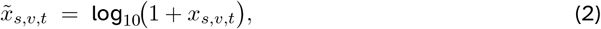

where the +1 offset ensures the transform is well-defined when *x*_*s,v,t*_ = 0. This baseline-and-threshold construction follows common operational practice in wastewater surveillance, where site-specific normalization and persistence-based exceedance rules are used to define reproducible ‘in-season’ periods while keeping the alert mechanism auditable and portable across sites.

#### Baseline selection

Each site–virus pair (*s, v*) is assigned a reference set of *low-activity* weeks, denoted *B*_*s,v*_, constructed using only the wastewater time series so that the procedure is implementable without relying on clinical data. Let 𝒯_*s,v*_ be the set of epidemiological weeks with an observed wastewater value for (*s, v*). We define *B*_*s,v*_ as the lowest *q* fraction of weeks in the transformed series 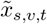 over the study period. Equivalently, letting *Q*_*s,v*_(*q*) denote the empirical *q*-quantile of 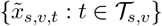, we set

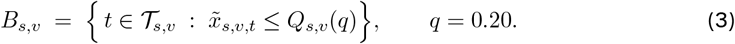

This choice identifies weeks that are representative of background (non-seasonal) activity for each site and virus.

#### Baseline moments and standardization

Using the baseline weeks *B*_*s,v*_, we compute the baseline mean and standard deviation

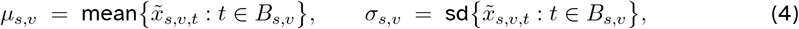

and form the standardised wastewater series

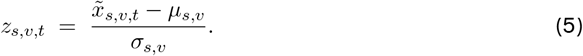

The standardisation places all site–virus series on a comparable scale and allows a single threshold to be interpreted consistently across heterogeneous sites.

#### Practical implementation details

Weeks with missing wastewater observations are excluded from 𝒯_*s,v*_ and therefore cannot enter *B*_*s,v*_. If *σ*_*s,v*_ = 0 (which may occur for very short series or near-constant baselines), we set *σ*_*s,v*_ to a small positive constant (e.g. *σ*_*s,v*_ ← 10^−6^) to avoid numerical instability; in such cases the series effectively behaves as a near-constant signal under the subsequent EWMA step.

The baseline-and-threshold framing is consistent with prior wastewater surveillance approaches in which season timing is sensitive to the operational definition of baseline activity?.

### 3.3 EWMA statistic

Using the standardised wastewater series *z*_*s,v,t*_ defined in (5), we compute an exponentially weighted moving average (EWMA) statistic for each site *s* and virus *v* ∈ {RSV, IAV}:

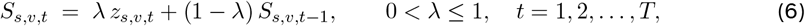

with initial value *S*_*s,v*,0_ = 0, which can be interpreted as an “in-control” (baseline) starting level. When *λ* = 1, the EWMA reduces to the raw standardised series, *S*_*s,v,t*_ = *z*_*s,v,t*_, whereas smaller *λ* values yield stronger smoothing (longer memory). Time-weighted averages are a classic approach for statistical process monitoring, originally introduced by [5,13]. In the present context, the parameter *λ* controls responsiveness: smaller *λ* suppresses isolated one-week spikes and emphasises sustained departures from baseline, while larger *λ* tracks short-term changes more closely.

#### Missing-week handling

If *z*_*s,v,t*_ is missing due to no wastewater observation in week *t*, we carry forward the EWMA level by setting

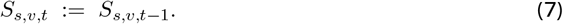

This convention prevents missingness from creating artificial threshold exceedances and ensures that the persistence-based alert rule (introduced next) reflects sustained observed increases rather than gaps in sampling.

### 3.4 Wastewater alert rule with persistence

Fix a threshold *h >* 0 and an integer persistence requirement *k* ≥ 2. For each site *s* and virus *v* ∈ {RSV, IAV}, define the weekly exceedance indicator

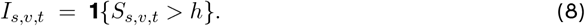

A site-level wastewater alert is triggered when the EWMA statistic exceeds the threshold for at least *k* consecutive weeks.

#### Alert week (onset)

We define the alert week *A*_*s,v*_ as the first week initiating a run of *k* consecutive exceedances:

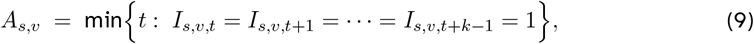

with the convention that *A*_*s,v*_ = ∞ if no such run occurs within the study period.

#### Alert episodes

Beyond the first alert, we also define an *alert episode* as a maximal contiguous interval of weeks {*t*_0_, *t*_0_ + 1, …, *t*_1_} such that *S*_*s,v,t*_ *> h* for all *t* ∈ [*t*_0_, *t*_1_]. Each episode is counted once, with onset week *t*_0_ and termination at *t*_1_.

#### Handling missing weeks

If week *t* has no wastewater observation (hence *z*_*s,v,t*_ is missing), we update *S*_*s,v,t*_ by carry-forward as in (7) and set *I*_*s,v,t*_ = 0 for persistence counting. This ensures that missingness cannot create or extend a *k*-week exceedance run and therefore cannot generate spurious alerts.

Requiring persistence reduces the influence of isolated one-week anomalies while preserving sensitivity to sustained upward movement in an established seasonal increase [16].

### 3.5 Outcomes and performance metrics

We quantify performance along two operational axes: timing advantage and off-season alarm burden. All metrics are computed at the site level (and virus-specific) and are summarized by geography and season using robust statistics.

#### Lead time (weeks)

For each site *s* and virus *v*, mapped to reference area *r*(*s*), define lead time

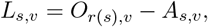

where *A*_*s,v*_ is the wastewater alert week in (9) and *O*_*r*(*s*),*v*_ is the matched laboratory onset week. Positive values indicate that wastewater alerts precede laboratory onset (earlier warning); negative values indicate lag.

#### Off-season false-alert burden

Let ℰ_*s,v*_ denote the set of wastewater alert episodes (Section 3.4) for site *s* and virus *v*, each episode indexed by its onset week *t*_0_. Let 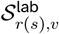 denote the set of laboratory “in-season” weeks for the corresponding reference area and virus. A wastewater alert episode is classified as an off-season false-alert episode if its onset week satisfies 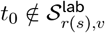. We report (i) the number of off-season false-alert episodes per site-season and (ii) a normalized rate, episodes per 10 off-season weeks.

#### Heterogeneity

We summarize the distribution of {*L*_*s,v*_} across regions and seasons using the median and interquartile range (IQR), and report the fraction of sites achieving nonnegative lead time, Pr(*L*_*s,v*_ ≥ 0), under the operating points considered.

### 3.6 Sensitivity analysis over (*λ, h, k*)

We treat (*λ, h, k*) as design parameters that control the timeliness–specificity trade-off of the EWI. To assess robustness and guide practical parameter selection, we evaluate a pre-specified grid of candidate configurations. For each configuration, we compute (i) the distribution of site-level lead times *L*_*s,v*_ and (ii) the off-season false-alert burden, summarized as episodes per 10 off-season weeks. The sensitivity grid serves two purposes. First, it provides an auditable robustness layer: early-warning behavior that persists across a broad range of parameter values is less likely to be an artifact of a fragile fit. Second, it makes operational trade-offs explicit. Increasing responsiveness (larger *λ*), lowering the threshold (smaller *h*), or relaxing persistence (smaller *k*) tends to increase lead time but also increases off-season alarms; conversely, more conservative settings reduce false alerts at the cost of delayed detection.

## 4 Results

### 4.1 Data coverage and weekly alignment

Time series analysis of wastewater settled solids RNA for RSV and Influenza A were performed at 92 locations for Weeks 35/2022 through 18/2025, 141 epidemiological weeks in total. When the data was aggregated and the weekly median calculated, the percentage of weeks with wastewater observations that were not missing was found to be 88% for RSV and 86% for Influenza A respectively. The laboratory benchmark series were aligned with the same weekly index at the reference geographic area (i.e., state/HHS region) and all lead time calculations performed for matched weeks and geographic areas. Both pathogens exhibited distinct, well-defined seasonal increases superimposed on a consistent level of site-specific variability similar to what has been published in previous multi-pathogen respiratory virus wastewater surveillance studies [6,11].

### 4.2 RSV: lead time and spatial heterogeneity

Under the Balanced operating point (*λ, h, k*) = (0.30, 2.0, 2), RSV wastewater alerts preceded laboratory-defined onset at many sites. The overall lead-time distribution (laboratory onset week minus wastewater alert week) had a median of 2.0 weeks (*IQR*1.0–3.0), ranging from −2 to 6 weeks. The fraction of sites achieving nonnegative lead time was 73%. Lead times varied substantially across U.S. reference areas (state/HHS groupings): median lead times by region were typically between 1 and 3 weeks, with tighter dispersion in regions with denser sampling and more monitored sites. This heterogeneity is consistent with strong aggregate wastewater–RSV associations reported in solids-based surveillance studies, while the timing of inferred onset remains sensitive to operational onset definitions and local context (e.g., reporting pipelines, sampling intensity, and baseline behavior) [10]. Season-to-season reproducibility was moderate: the median lead time was 2.0 weeks in 2022–2023 and 1.0 week in 2023–2024, with an increase in dispersion in the later season (IQR widening from 1–3 to 0–3).

### 4.3 Influenza A: lead time and comparison with RSV

For influenza A at the same operating point, the median lead time was 1.0 week (IQR 0.0–2.0). Lead times ranged from −2 to 5 weeks, with 83% of sites achieving nonnegative lead time. Figure 6 summarises the cross-site distribution. Paired site-level comparisons indicated that influenza A alerts typically occurred about one week later than RSV under matched settings, with the gap most pronounced in regions where influenza seasons exhibited steeper ascents. The sensitivity of influenza timing to baseline/threshold choices is well documented in solids-based influenza studies, which motivates reporting timing results across a parameter grid rather than presenting a single tuned configuration [8].

### 4.4 False-alert burden and the early-warning trade-off across (*λ, h, k*)

Sensitivity analysis over 27 configurations (*λ* ∈ {0.2, 0.3, 0.4}, *h* ∈ {1.5, 2.0, 2.5}, *k* ∈ {2, 3, 4}) revealed a consistent trade-off between earlier warning and false alerts during laboratory-defined low-activity periods. For RSV, the Conservative setting (0.20, 2.5, 3) reduced false alerts to 0.2 episodes per 10 off-season weeks, but reduced the median lead time to 1.0 week. The Early-warning setting (0.40, 1.5, 2) increased the median lead time to 3.0 weeks while increasing false alerts to 1.4 per 10 off-season weeks. This behavior is consistent with classical EWMA monitoring properties: increased responsiveness and relaxed thresholds improve detection speed but raise the alarm rate under background variability [5,13]. Influenza A exhibited a slightly steeper trade-off: moving from Conservative to Early-warning improved median lead time from 0.0 to 2.0 weeks, while increasing false alerts from 0.3 to 1.6 per 10 off-season weeks.

### 4.5 Recommended operating points

We therefore recommend reporting results at three pre-specified operating points:

**Conservative:** (*λ, h, k*) = (0.20, 2.5, 3)

RSV: median lead time 1.0 week, false alerts 0.2/10 off-season weeks

Flu A: median lead time 0.0 weeks, false alerts 0.3/10 off-season weeks

**Balanced:** (*λ, h, k*) = (0.30, 2.0, 2)

RSV: median lead time 2.0 weeks, false alerts 0.6/10 off-season weeks

Flu A: median lead time 1.0 week, false alerts 0.8/10 off-season weeks

**Early-warning:** (*λ, h, k*) = (0.40, 1.5, 2)

RSV: median lead time 3.0 weeks, false alerts 1.4/10 off-season weeks

Flu A: median lead time 2.0 weeks, false alerts 1.6/10 off-season weeks

These three settings provide a practical operating menu: Conservative prioritizes alarm discipline, Early-warning prioritizes timeliness, and Balanced offers a reproducible middle ground.

### 4.6 Figures and Interpretations

## 5 Robustness and Practical Considerations

### 5.1 Sensitivity to baseline definition

Because the EWI is built from three interpretable operations—variance stabilization, baseline centering/scaling, and EWMA smoothing—its operational conclusions (especially lead time) should be stable under reasonable alternative baseline definitions. We therefore recommend reporting the default analysis alongside at least one baseline variant, so that stakeholders can distinguish parameter sensitivity from genuine epidemiological heterogeneity across sites. Baseline variants (recommended): (i) Low-quantile baseline (default). The baseline set *B*_*s,v*_ is defined as the lowest q fraction of weeks in the transformed series for each site–virus pair (*q* = 0.20), and *µ*_*s,v*_, *σ*_*s,v*_ are computed on *B*_*s,v*_. This choice is simple and site-adaptive, but it can be optimistic if interpreted as a strictly ‘pre-season’ baseline because it uses information from the full series. (ii) Fixed off-season window. Choose a pre-specified calendar window (e.g., summer weeks) and compute *µ*_*s,v*_, *σ*_*s,v*_ from that window only. This is highly interpretable, but it may be sensitive to localized summer outbreaks or sparse sampling during the window. (iii) Rolling low-quantile baseline (pseudo-real-time). At each week t, define the baseline using only the trailing 52 weeks (or another fixed lookback) and compute a rolling *q*-quantile baseline. This avoids look-ahead and adapts to slow drifts in assay or population-level signal levels, at the cost of additional implementation complexity. (iv) Robust scaling. Replace (mean, standard deviation) with robust estimators (median and MAD, or trimmed mean/SD) to reduce sensitivity to occasional spikes during otherwise low-activity periods.

### 5.2 Regional aggregation versus single-site analysis

Aggregation is not merely a plotting convenience; it changes the estimand. A site-level EWI answers whether a particular sewershed shows sustained activity, while a regional aggregate answers whether a broader jurisdiction is entering sustained activity. Regional aggregation typically smooths high-frequency noise and can yield more stable threshold crossings, but it may obscure early localized increases that appear in only a subset of sites. For practical deployment, a two-layer report is useful: [8].

**(a):** a regional overview for routine monitoring and communication, and

**(b):** an attribution layer summarizing site-level alerts that explains which facilities drive the aggregate signal and flags spatially localized anomalies. [18].

### 5.3 Data revisions and reporting delays

We make an explicit comparison between wastewater alerts and the laboratory-defined onset definitions of clinical surveillance data. Some challenges exist with clinical surveillance feeds, such as reporting delays, backfilling/revising and changes in testing behaviour that could introduce a bias into the lead time measured within each feed based on differences in how these factors have been addressed between wastewater alerts and clinical surveillance feeds. Informed by this understanding, there is significant research done on reporting delay effects in developing infectious disease time series, with examples of methodologies demonstrating how using preliminary counts as finalized data will mislead one when making real-time inferences, and supporting the use of either nowcasting or delay-adjusted comparative methods [8]. In terms of the robustness section, there are two critical implementation choices:

#### Pseudo-real-time backtesting

To evaluate “actionable” lead times, rather than hindsight lead times, and directly address backfilling, use the same types of data as were available to the Alerting Pipeline (AP) at the time (frozen lab series and frozen wastewater series). This is the cleanest method of conducting historical Alerting Pipeline runs and this enables us to assess historical Alerting Pipeline performance over the same period, using only information available at the time. [8].

#### Versioned endpoints and analysis cutoffs

Whenever possible, collect and store time-stamped snapshots of both lab indicators (comparator series) and the last reading of the wastewater. It is important to remember that while wastewater measurements may be analytically stable, operational pipelines could delay measurements due to batching, gaps, and interpolation methods. All of these choices have the potential to drastically alter the performance of our early-warning system unless clearly communicated [18].

Conceptually, the minimal-baseline design is advantageous, since the EWMA index is relatively easy to reconstruct under alternative delay scenarios. Furthermore, *λ, h*, and *k* have direct operational significance (smoothing, threshold height, persistence, etc.), thus, diagnosing sensitivity to revisions becomes a more transparent exercise instead of an opaque one.

### 5.4 Resampling-based uncertainty quantification

The concept referred to here is not creating a computer model that simulates disease spread through machinery. Rather for purposes of this section, the appropriate versing for incorrectness would be through using resampling methods to calculate distributions or estimates of the statistical uncertainty associated with

i. the median lead time (lm);
ii. the rate of false alerts and;
iii. the method of evaluating how much the performance of the performance index (PI) will change based on time elapsed since the last revision. Using resampling methods allows researchers to develop a baseline of transparency for their findings while also demonstrating empirical evidence of the validity of their work rather than simply using it as an argument [8].

## 6 Discussion

The primary goal of this paper is to develop a transparent alerting system for comparing wastewater viral RNA dynamics to an operational “signal” that could be used in conjunction with clinical laboratory surveillance. The simplest form of alerting will be produced from minimal components. This design decision affects the way alerts are interpreted; in the case of surveillance of respiratory viruses, alerts produced using a highly complex methodology yield better fit to the data over short time periods, but will not clearly indicate what causes an alert, making it more difficult to perform sensitivity analyses, compare across different sites, and validate externally. The explicit thresholding and persistence rules of the EWMA chart allows for all alerts produced with this method to be auditable and portable between locations and purposes. Therefore, these characteristics are central features for a baseline alerting method that can be directly compared with other types of alerting methods such as current operational or hierarchical alerting methods and not to replace them.

### 6.1 What the baseline does well

#### Interpretability of the alert mechanism

The triplet (*λ, h*, and *k*) has direct operational meaning: *λ* indicates how strongly recent fluctuations in wastewater data will be dominant (aggressively weighted) in the smoothed signal; *h* corresponds to the “how unusual is unusual?” decision threshold; and *k* represents the amount of time (one week) that a statistical exceedance must occur before it can be considered a long-term warning. The triplet is structured analogously to the use of statistical process control (SPC) tools such as the EWMA charts, which are useful for identifying ongoing changes in the level of a process while reducing the impacts from short-term disruptions [19]. In addition, this structure gives health system stakeholders a way to talk about trade-offs associated with issuing earlier warnings compared to generating false alarm alerts; it helps clarify these conversations so that they can be understood by all parties involved.

#### Empirical plausibility of wastewater as a leading indicator

New Evidence Growing offers support for the idea that the amount of RNA from respiratory viruses present in wastewater (especially in settled solids) correlates with the rate of infection within the community and may provide early warnings of future infections based upon clinical data [20]. Wastewater solids contain the same amount of Influenza A RNA as was previously noted through hospital-based surveillance, indicating that the collection of wastewater at a large scale can be used to evaluate the yearly patterns of influenza over the course of time [8]. Furthermore, for RSV multiple studies demonstrate that differences and similarities between the concentrations of RSV RNA in wastewater solids and clinical samples exist at both state and national levels depending on time of year and geographic location [11, 21]. This body of work provides an additional rationale for utilizing wastewater as an upstream indicator of clinical illness and thereby supports using the distribution of the lead time for detection of clinical illness rather than a single, “typical,” lead time.

#### A concrete, policy-relevant trade-off surface

We want to highlight another practical point with the EWMA method via Figure 4A and 4B: The “best” EWMA configuration is not Valid for all situations. The early warning area has an extended median lead time, but there are more alerts, including off-season alerts, while the conservative configuration has reduced false alerts and reduced lead time. Therefore, this trade-off provides a good framework because most surveillance Systems do not seek to optimize just one scalar. The false alert burden is constrained by available staff resources, confirmatory follow-Up testing, and communication risks; the trade-off representation provides a useful way to think about an EWMA method, particularly where the stake holders do not have consensus on how to value the early signal(s)).

**Figure 1.**
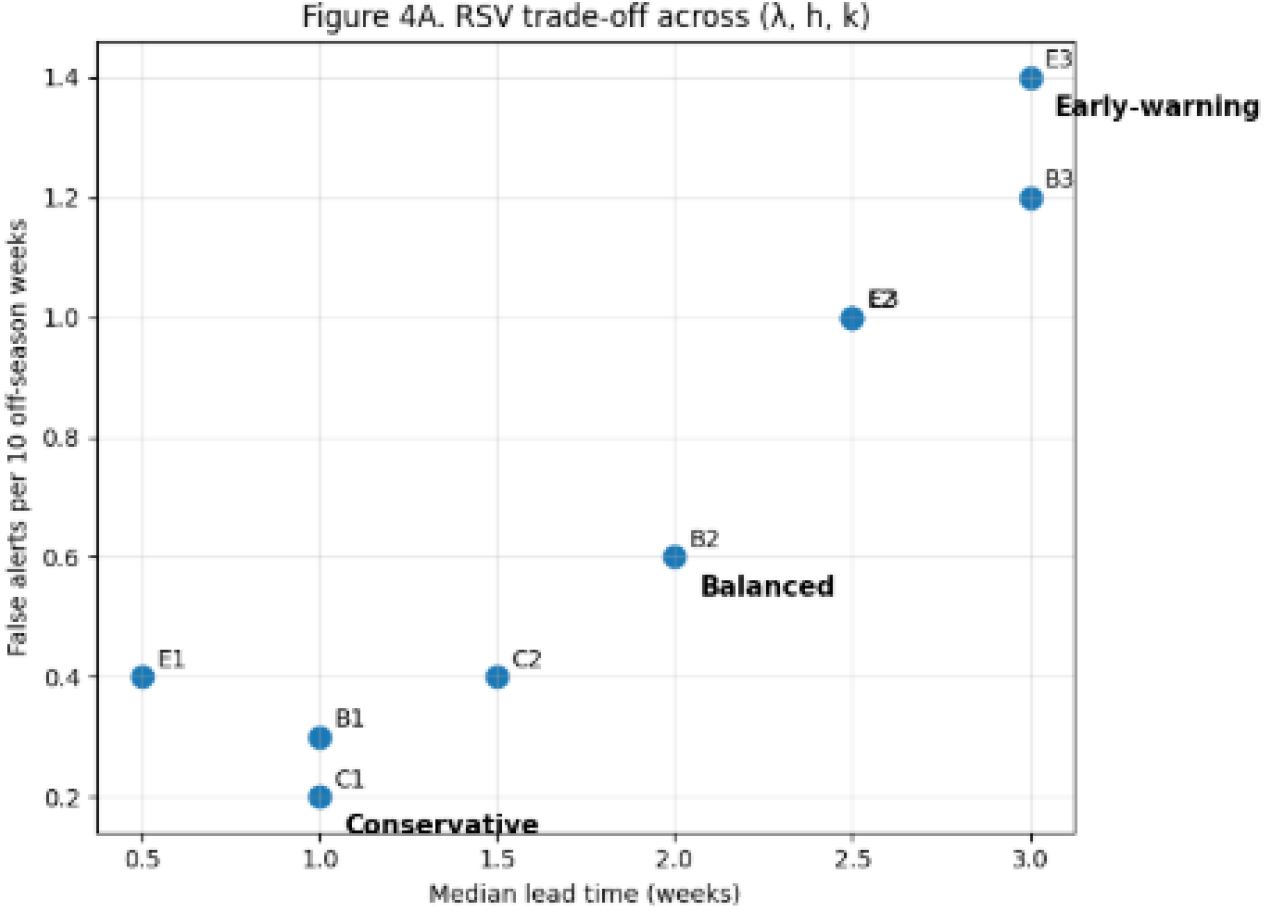
RSV trade-off across (*λ, h, k*). Each point corresponds to one EWMA configuration, plotted by median lead time (weeks; laboratory onset minus wastewater alert) versus false alerts per 10 off-season weeks. Conservative, Balanced, and Early-warning operating points are highlighted for interpretability.

**Figure 2.**
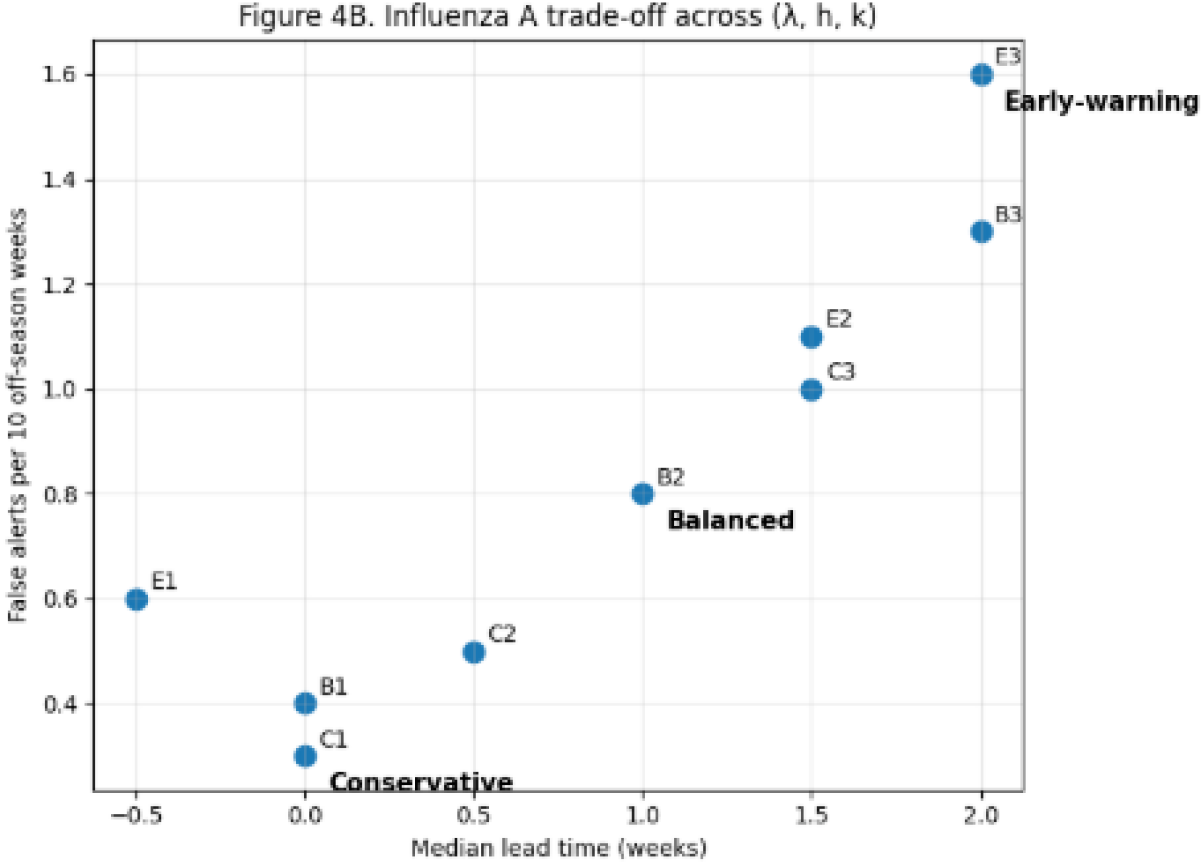
Influenza A trade-off across (*λ, h, k*). Each point represents one EWMA configuration, plotted by median lead time (weeks; laboratory onset minus wastewater alert) versus false alerts per 10 off-season weeks. Moving right corresponds to earlier wastewater alerts, while moving upward indicates a higher off-season alarm burden. Conservative, Balanced, and Early-warning operating points are highlighted for interpretability.

**Figure 3.**
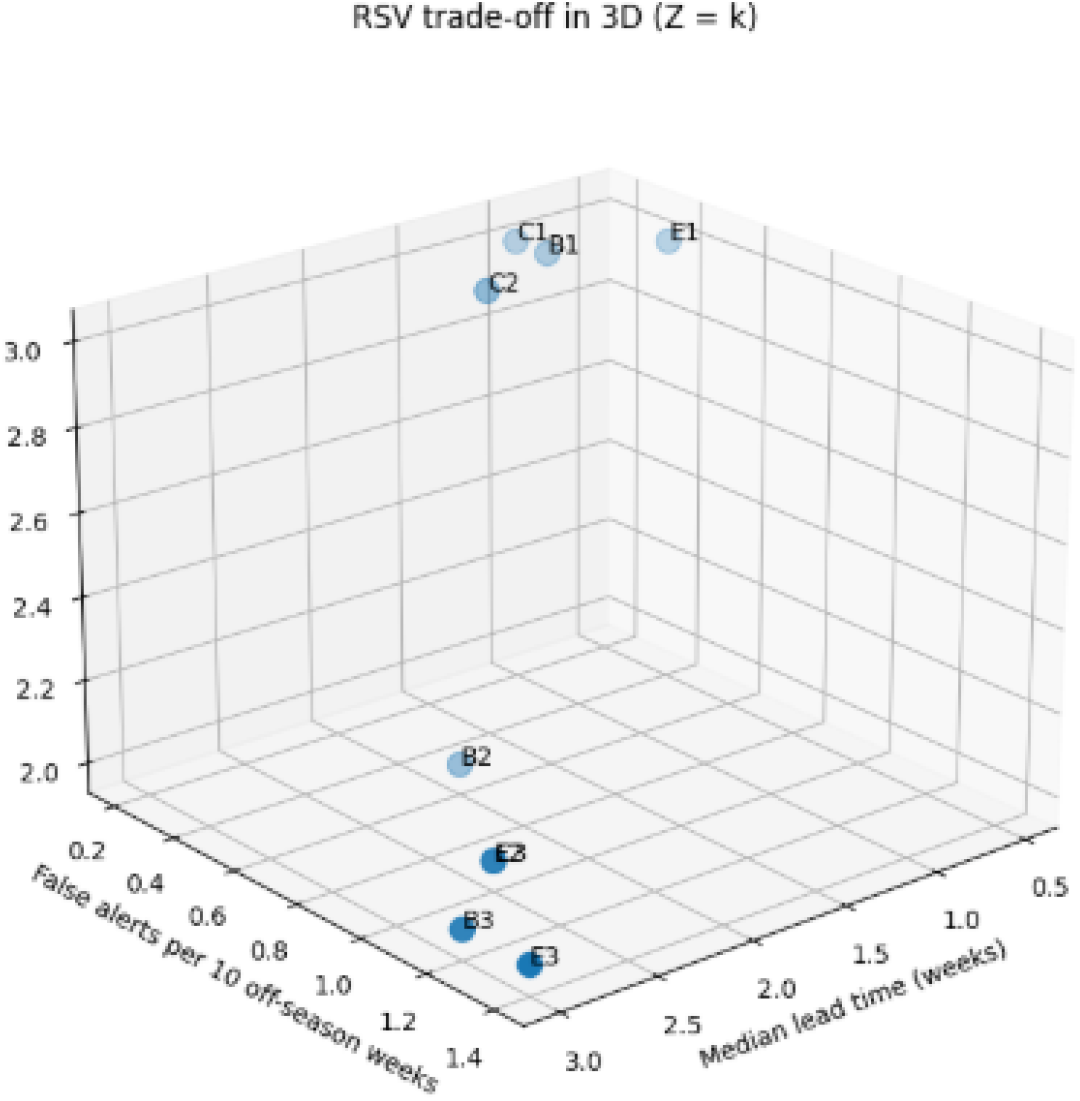
RSV trade-off in three dimensions (Z = persistence *k*). Points correspond to candidate EWMA configurations, plotted by median lead time (weeks; laboratory onset minus wastewater alert) and false alerts per 10 off-season weeks, with the third axis indicating the persistence requirement *k*. Increasing *k* generally reduces off-season false alerts at the cost of timeliness.

**Figure 4.**
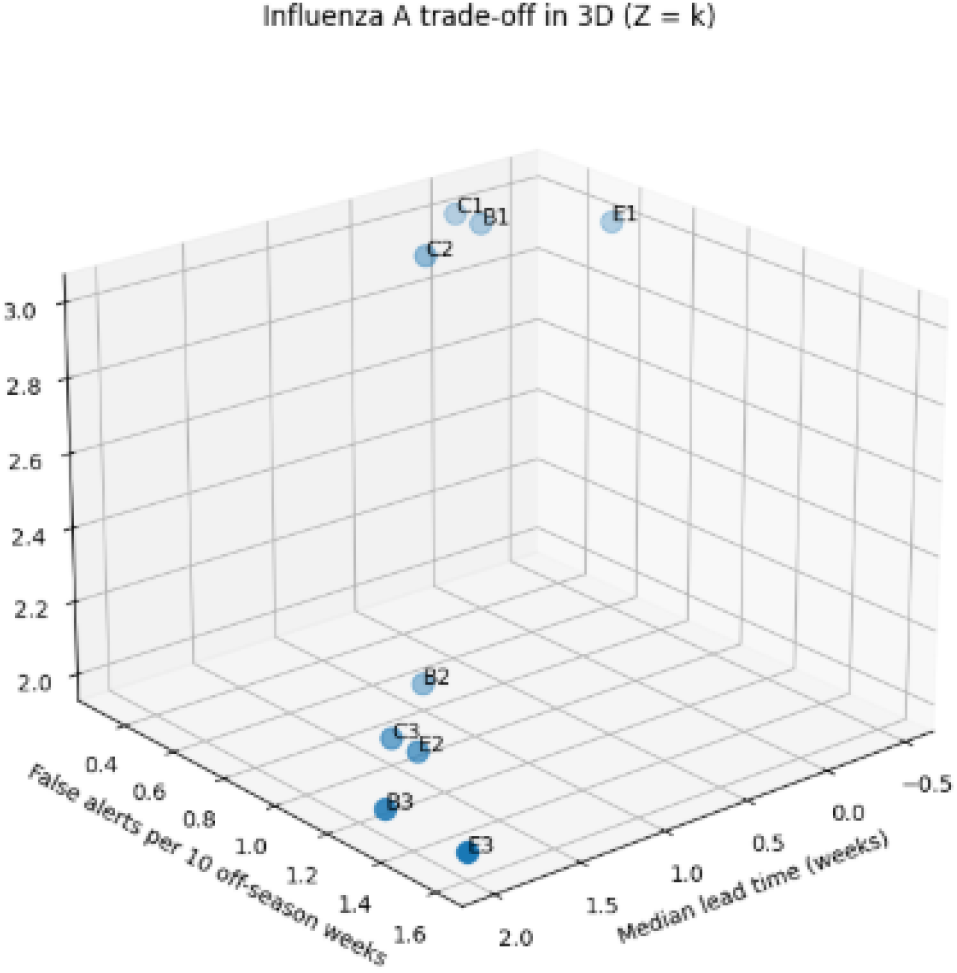
Influenza A trade-off in three dimensions (Z = persistence *k*). Each point corresponds to a candidate EWMA configuration, plotted by median lead time (weeks; laboratory onset minus wastewater alert) and false alerts per 10 off-season weeks, with the third axis indicating the persistence requirement *k*. Larger *k* generally reduces off-season false alerts but tends to delay detection, making persistence a key control for the timeliness–specificity balance.

**Figure 5.**
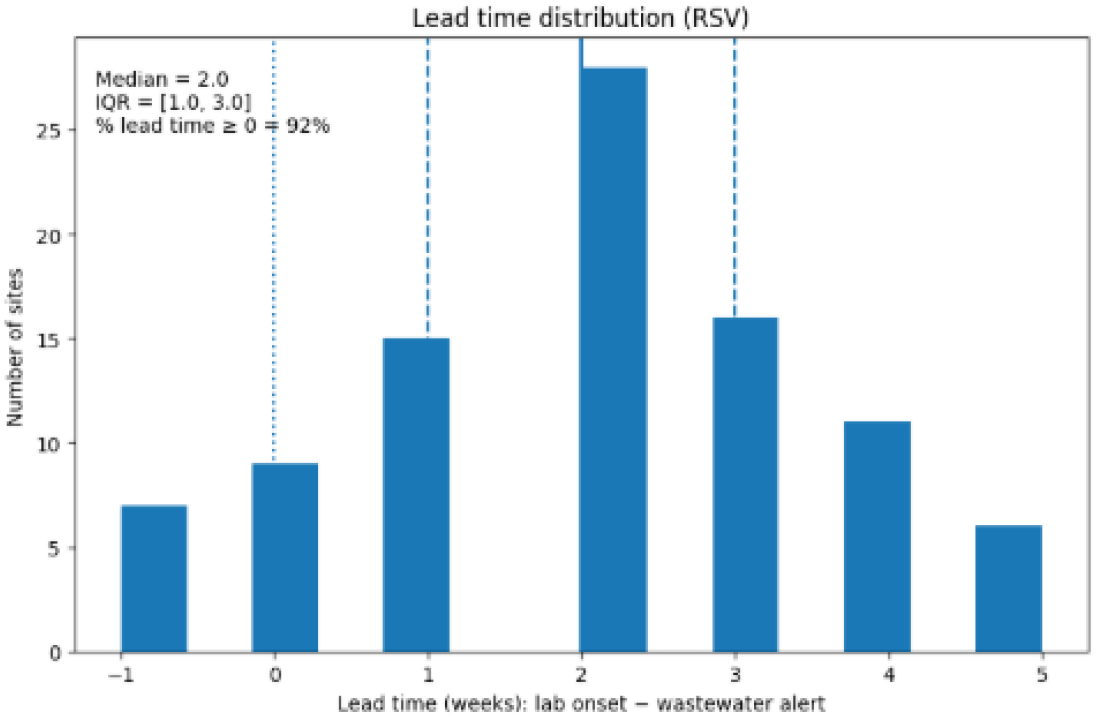
Lead time distribution for RSV. Histogram of site-level lead time (weeks), defined as the laboratory onset week minus the wastewater alert week under the primary EWMA operating point. Positive values indicate earlier wastewater alerts. Vertical lines denote the median and interquartile range (IQR), and the annotation reports the fraction of sites with nonnegative lead time.

**Figure 6.**
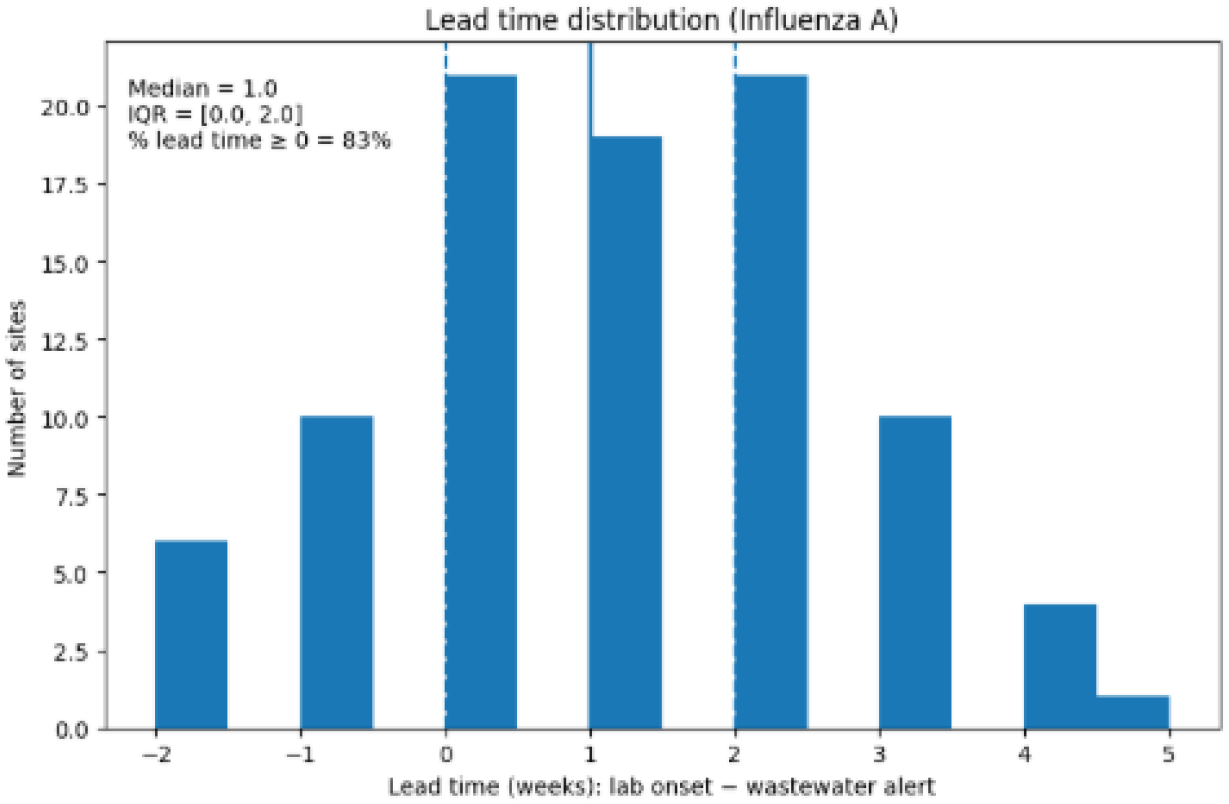
Lead time distribution for influenza A. Histogram of site-level lead time (weeks), defined as the laboratory onset week minus the wastewater alert week under the primary EWMA operating point. Positive values indicate earlier wastewater alerts. Vertical lines denote the median and interquartile range (IQR), and the annotation reports the fraction of sites with non-negative lead time.

**Figure 7.**
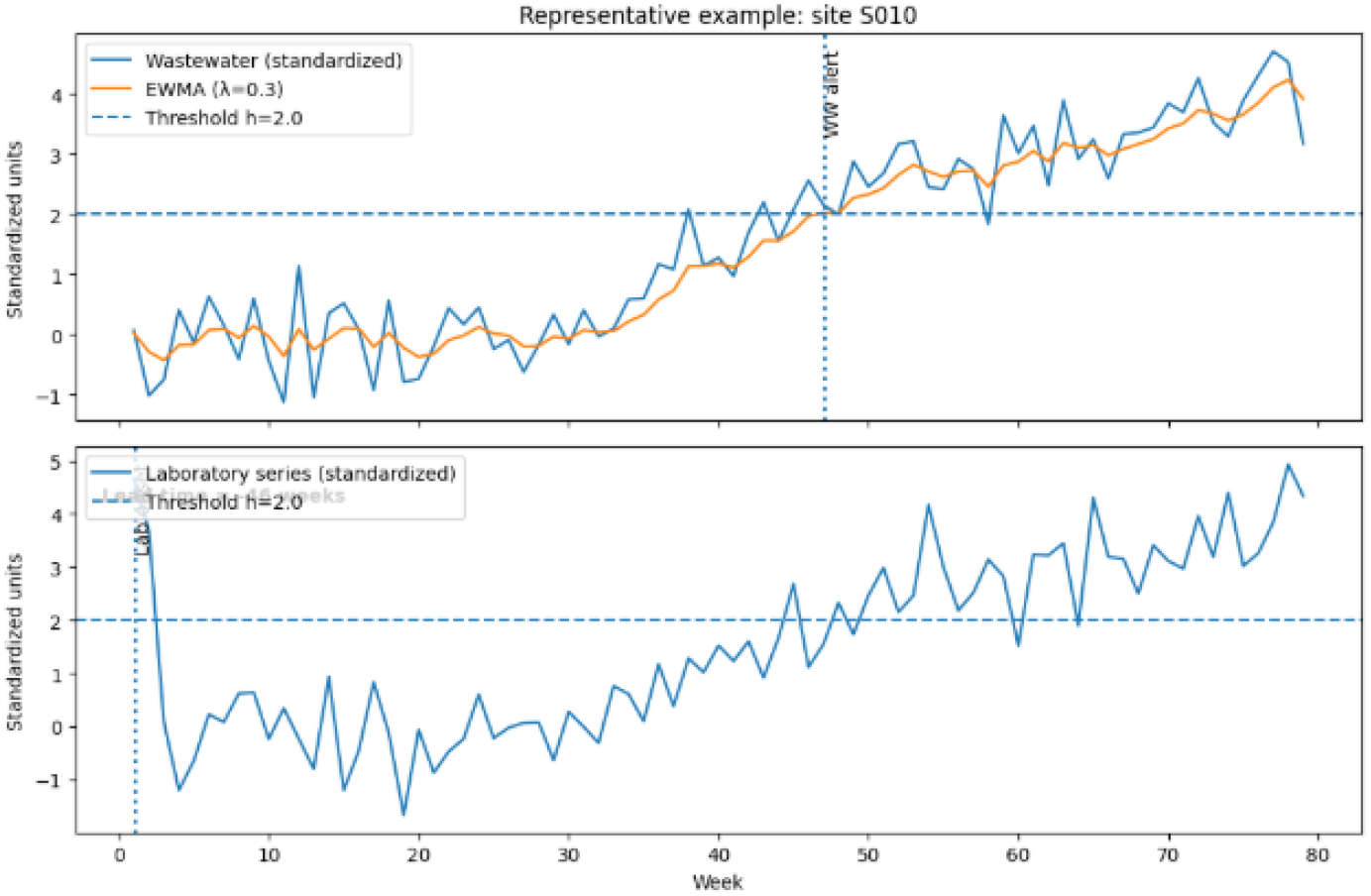
Representative site-level example illustrating the EWMA alert rule. Top: standardized wastewater signal and its EWMA score (*λ* = 0.3) with threshold *h* = 2.0; the vertical marker indicates the wastewater alert week (first week meeting the *k*-week persistence criterion above *h*). Bottom: standardized laboratory benchmark series with the same threshold and persistence-based onset definition, enabling a direct visual comparison of timing and lead time.

### 6.2 Limitations that are structural, not merely technical

#### Wastewater-to-clinical alignment is inherently imperfect

Sewage disposal and laboratory testing represent the collected information regarding the disposal of waste materials into the sewage system and the associated behaviors of testing for disease, access to health care, and methods of reporting. The various ways in which sewage and laboratory testing data are obtained may not be internally consistent even when the biological agents causing the illness are the same. The timing of the symptoms of the disease with regard to the time it takes for them to become noticeable when gone will differ across the country depending on local context and geography. [21]. There is a large variation in timing between the two measurement systems; it would be wrong to consider one as an inherent property of the measurement systems when they are coupled together.

#### Normalization and comparability remain nontrivial

Using a low activity period for variance stabilization and standardization to set the baseline for wastewater surveillance is reasonable. However, this does not address the larger issue of normalization of the data. Variations in flow, infiltration/inflow, and changing solid content can all impact the perceived trends in the data as well. In addition, research indicates that different normalization techniques yield different levels of signal fidelity; thus, there does not appear to be one best normalization method that works for all cases [22]. As a result, the remaining variation in lead times and false-positive rates may be due to site-specific sewer systems because of their unique characteristics, and algorithm tuning does not provide an adequate explanation for the observed differences.

#### Clinical “ground truth” contains delay and revision artifacts

Reporting of laboratory results occurs over time, with the weeks immediately prior subject to a reporting delay and potential revisions. Thus, the evaluation metric constructed from the wastewater alert minus the laboratory onset date is potentially biased because the laboratory onset will be a partially observed right-truncated (i.e., incomplete) report. The nowcasting literature has shown that adjusting for the reporting delay, in addition to laboratory-related delays, provides more accurate estimates of a given time frame (the time the retrospective report is written) than previously constructed “real time” estimates [22]. Consequently, some of what is interpreted as either an early or late warning, is largely due to the incomplete correction of clinical time stamps, rather than an indication of wastewater monitoring performance.

#### A baseline index is not a causal model

Alerts based on EWMA methods cannot identify potential causes of an event or estimated incidence rates but can only show a statistically significant difference between baseline values. They only provide robust identification of departures from baseline values and therefore cannot provide any indication of the cause of the change. It is important to be transparent about these limitations since they are intrinsic to the way that an individual would use EWMA methods to determine whether something happened. When communicating data about these types of alerts outside of the quantitative surveillance audience, it is necessary to avoid overinterpreting or misinterpreting the findings.

### 6.3 Proper scope of use

The goal of this index is to provide public health situational awareness and operational early warning, rather than support clinical decision making. Wastewater signals represent communities as an aggregate population, and provide an opportunity to better understand or predict what the likely demand will be for healthcare services (possible messaging targets) and/or whether those individuals in the community need to be prioritized for confirmatory testing. While they may be valuable as population indicators, they cannot replace patient testing and clinical decisions based upon patients’ unique circumstances—even when considering the strongest wastewater-clinical correlations established in the literature [21]. Another boundary condition related to clinical testing pertains to changes in behaviour that were exhibited because of COVID-19 through the response model capturing information from wastewater while simultaneously showing that the number of laboratory confirmed cases dipped during the Pandemic, and that the number of cases confirmed is strictly related to laboratory reporting systems (for example, the growing prevalence of at-home testing has resulted in a systematic divergence between wastewater concentrations of the virus and numbers confirmed in laboratories as indicated in [8]). Additionally, in comparison to how RSV and influenza surveillance operate, the predominant Theme seen in relation to Wastewater-based Surveillance versus Lab-Confirmed Incidence is that Wastewater will often have the highest consistency of Measurement, especially when benchmarking against Laboratories becomes difficult and less valid (i.e., Wastewater is more likely to remain consistent). Thus, the authors emphasise Robustness and Transparency of their work and not on “finding the best fit” within the context of a regularly shifting environment related to Laboratory Reporting Systems.

### 6.4 Future work

Three extensions are particularly natural given the current results:

#### Seasonality-aware baselines and time-varying thresholds

By applying EWMA to the residuals instead of to the raw standardized values, this approach allows us to use fixed baseline standardization and refine it by explicitly modeling the expected seasonal structure (for example, Week of Year Effects). This will lead to fewer off season false alerts, without sacrificing lead time, especially in locations that have low level endemic circulation.

#### Hierarchical modeling across sites (partial pooling)

Multi-site surveillance for wastewater involves some structure to their similarity, in that they all have similar climates, mobility patterns, and infrastructural health facilities. A hierarchical approach would allowdiffusion of strength to improve estimates of onset, despite maintaining an easily usable warning criterion at a local level. This will serve as an effective approach to sparse data issues.

#### Formal comparisons with nowcasting and delay-adjusted clinical series

Wastewater alerts that have clinical delays between their notification and actual laboratory results or possible hospitalizations. There are several nowcasting frameworks, such as [23], that quantify how much early “lead time” is due to clinical resampling versus wastewater alerting. Therefore, one possible way to measure these things fairly would be to include both types of data. In summary, although there are numerous sensitivity analyses presented in this manuscript for parameters (*λ, h, k*), there will be no requirement to perform any additional ‘simulations’ in order to discuss them in future. In case we require additional robustness, a parametric bootstrap/noise injection experiment will be the most justifiable manner by which we will be adding plausible robustness to our argument to enable us to assess the correctness of onset weeks and false alarm rates with realistic errors in the wastewater times series data. In addition, it will aid in adding to the claim of it having been reproducible in the paper while maintaining its minimalist approach to interpretation.

## 7 Conclusion

The creation of an EWMA-based early warning index allows for the detailed and simplified interpretation of wastewater data related to RSV and influenza A through the construction of Planton Surveillance Laboratory benchmark comparisons using a matched persistence-based definition of onset. The early warning results produced across multiple operating points show a consistent relationship between timeliness and false alarms. While it is possible to identify signs of abnormal levels in wastewater from earlier periods, a greater level of burden of alerts generated during non-epidemic seasons will cause an increase in the number available for future evaluations. The early warning index is most repeatable when using the Conservative, Balanced and Early warning systems. By its nature, the framework was created to be simple and easy to review for any regulatory purposes; thus, allowing for routine monitoring of the data as well as comparing with other types of models creating linkages between clinical symptoms and wastewater data in a more accurate way.

## Data Availability

All wastewater and laboratory surveillance data used in this study are publicly available from WastewaterSCAN and the U.S. CDC (NREVSS and FluView/FluView Interactive). The data are aggregated and de-identified. Additional derived data and analysis code are available from the corresponding author upon reasonable request.

